# Diversity and Utilization Patterns of Medicinal Plants Used in the Management of Diabetes Mellitus: An Ethnobotanical Study in Selected Communities in Sierra Leone

**DOI:** 10.64898/2026.07.18.26358386

**Authors:** Sallu Nfagie Kamara, Abraham Isiaka Jimmy, Lee Presley Gary

## Abstract

**Background:** Diabetes mellitus is an increasing public health challenge in Sierra Leone, where access to diagnosis, treatment, and long-term care remains limited. Traditional medicine continues to play a significant role in disease management; however, ethnobotanical knowledge related to diabetes remains insufficiently documented.

**Methods:** A cross-sectional ethnobotanical survey was conducted among 40 informants, including traditional healers, herbalists, and knowledgeable community members in Waterloo, Pendembu, and Bo. Data were collected using structured questionnaires administered via Kobo Toolbox and paper-based tools. Information on medicinal plants, plant parts used, preparation methods, routes of administration, and knowledge transmission pathways was obtained. Quantitative ethnobotanical indices, including Frequency of Citation (FC), Relative Frequency of Citation (RFC), and Informant Consensus Factor (ICF), were calculated.

**Results:** A total of 21 medicinal plant species were documented. The most frequently cited species were *Moringa oleifera* (FC = 9; RFC = 0.225), *Vernonia amygdalina* (FC = 7; RFC = 0.175), and both *Cassia siberiana* and *Telfairia occidentalis* (FC = 6; RFC = 0.150). Leaves were the most commonly utilized plant part (40.9%), and decoction was the predominant preparation method (76.2%), with oral administration accounting for 95.2% of use. The Informant Consensus Factor (ICF = 0.69) indicated a relatively high level of agreement among informants. Knowledge was primarily transmitted through apprenticeship and inherited family practices.

**Conclusion:** Traditional medicinal plants remain an important component of diabetes management in Sierra Leone. The high level of consensus among informants and the repeated citation of specific plant species suggest structured and culturally validated therapeutic practices. The findings provide a foundation for future phytochemical and pharmacological investigations and highlight the need for documentation, preservation, and sustainable utilization of ethnobotanical knowledge.

## 1. Introduction

Diabetes mellitus is a chronic metabolic disorder characterized by persistent hyperglycaemia resulting from defects in insulin secretion, insulin action, or both. It is associated with disturbances in carbohydrate, lipid, and protein metabolism and leads to long-term complications affecting multiple organs, including the kidneys, eyes, nerves, and cardiovascular system (World Health Organization, 2024). The global burden of diabetes has increased significantly over the past few decades, making it one of the most pressing public health challenges of the 21st century.

According to the International Diabetes Federation (IDF), approximately 537 million adults were living with diabetes in 2021, with projections suggesting that this number may rise to 783 million by 2045 (International Diabetes Federation, 2024; NCD-RisC, 2024). More recent estimates from the Noncommunicable Disease Risk Factor Collaboration indicate even higher figures, reaching up to 828 million globally (NCD-RisC, 2024). Alarmingly, nearly half of individuals living with diabetes remain undiagnosed, particularly in low- and middle-income countries, where healthcare systems are often under-resourced.

Sub-Saharan Africa is among the regions experiencing the fastest increase in diabetes prevalence, driven by rapid urbanization, lifestyle changes, dietary transitions, and aging populations (Hossain et al., 2024; Kassa et al., 2024). In Sierra Leone, emerging evidence suggests a growing burden of diabetes, with increasing cases reported in both urban and rural settings (Kamara et al., 2024). However, access to diagnostic services, trained healthcare personnel, and essential medicines remains limited, contributing to poor disease management and increased risk of complications.

In this context, traditional medicine continues to play a critical role in healthcare delivery. It is estimated that up to 80% of populations in developing countries rely on traditional medicine for primary healthcare needs (Oyebode et al., 2016; World Health Organization, 2023). Traditional healing systems incorporate the use of medicinal plants, polyherbal formulations, and culturally embedded therapeutic practices. In Sierra Leone, herbal medicine remains widely used, particularly in rural and peri-urban communities where access to formal healthcare is limited (Vandy et al., 2026).

Ethnobotanical research has demonstrated that many medicinal plants possess bioactive compounds with antidiabetic properties, including flavonoids, alkaloids, terpenoids, and phenolic compounds. These phytochemicals exert hypoglycaemic effects through mechanisms such as enhancement of insulin secretion, improvement of insulin sensitivity, inhibition of carbohydrate-digesting enzymes, and antioxidant activity (Alam et al., 2022; Chahrour et al., 2025).

Several studies across Africa have documented medicinal plants used in diabetes management. For example, ethnobotanical surveys in Uganda, Ethiopia, and the Democratic Republic of Congo have reported a wide diversity of plant species used traditionally for glycaemic control (Amuri et al., 2018; Tugume & Nyakoojo, 2019; Agize et al., 2022). These studies highlight the importance of indigenous knowledge systems in identifying potential therapeutic agents.

Despite this growing body of evidence, systematic documentation of medicinal plants used for diabetes management in Sierra Leone remains limited. Existing studies are often localized and lack comprehensive analysis of plant diversity, preparation methods, and knowledge transmission pathways (Adepoju et al., 2023). This gap limits opportunities for pharmacological validation, conservation planning, and integration of traditional medicine into national healthcare systems.

Therefore, this study aims to document and analyze medicinal plants used in the management of diabetes mellitus in selected communities in Sierra Leone, while also examining patterns of plant use, preparation methods, and knowledge transmission. By applying quantitative ethnobotanical indices, this research provides a structured approach to identifying culturally significant plant species and generating evidence for future scientific investigation.

## 2. Materials and Methods

### 2.1 Study Design

A cross-sectional ethnobotanical survey was conducted to document medicinal plants used in diabetes management.

### 2.2 Study Area

The study was conducted in three communities in Sierra Leone:

- Waterloo (Western Area)
- Pendembu (Eastern Province)
- Bo (Southern Province)

### 2.3 Study Population

A total of 40 informants, including traditional healers, herbalists, and knowledgeable community members, were purposively selected.

### 2.4 Data Collection

Data were collected using structured questionnaires administered via Kobo Toolbox and paper-based tools. Information collected included:

- Plant species used
- Plant parts utilized
- Preparation methods
- Routes of administration
- Knowledge transmission pathways

### 2.5 Data Analysis

Ethnobotanical surveys are widely recognized as effective tools for documenting indigenous medicinal knowledge and identifying plant species with potential pharmacological relevance (Tugume & Nyakoojo, 2019; Ong et al., 2020). The use of quantitative indices such as Frequency of Citation (FC), Relative Frequency of Citation (RFC), Informant Consensus Factor (ICF), and Fidelity Level (FL) enhances the reliability of ethnobotanical data by providing objective measures of cultural importance and agreement among informants (Silva et al., 2017; Ong et al., 2020). These indices are commonly applied in ethnopharmacological research to prioritize plant species for further phytochemical and pharmacological investigation.

Ethnobotanical indices were calculated:

- **Frequency of Citation (FC)**
- **Relative Frequency of Citation (RFC = FC/N)**
- **Informant Consensus Factor (ICF)**

Descriptive statistics were used to summarize the data.

## 3. Results

### 3.1 Socio-Demographic Characteristics of Respondents

A total of 40 informants participated in the ethnobotanical survey conducted across the selected communities. The demographic characteristics of respondents are summarized according to age distribution, sex, marital status, educational attainment, occupation, ethnicity, religion, community role, and experience in traditional medicinal practices.

**Table 1.**
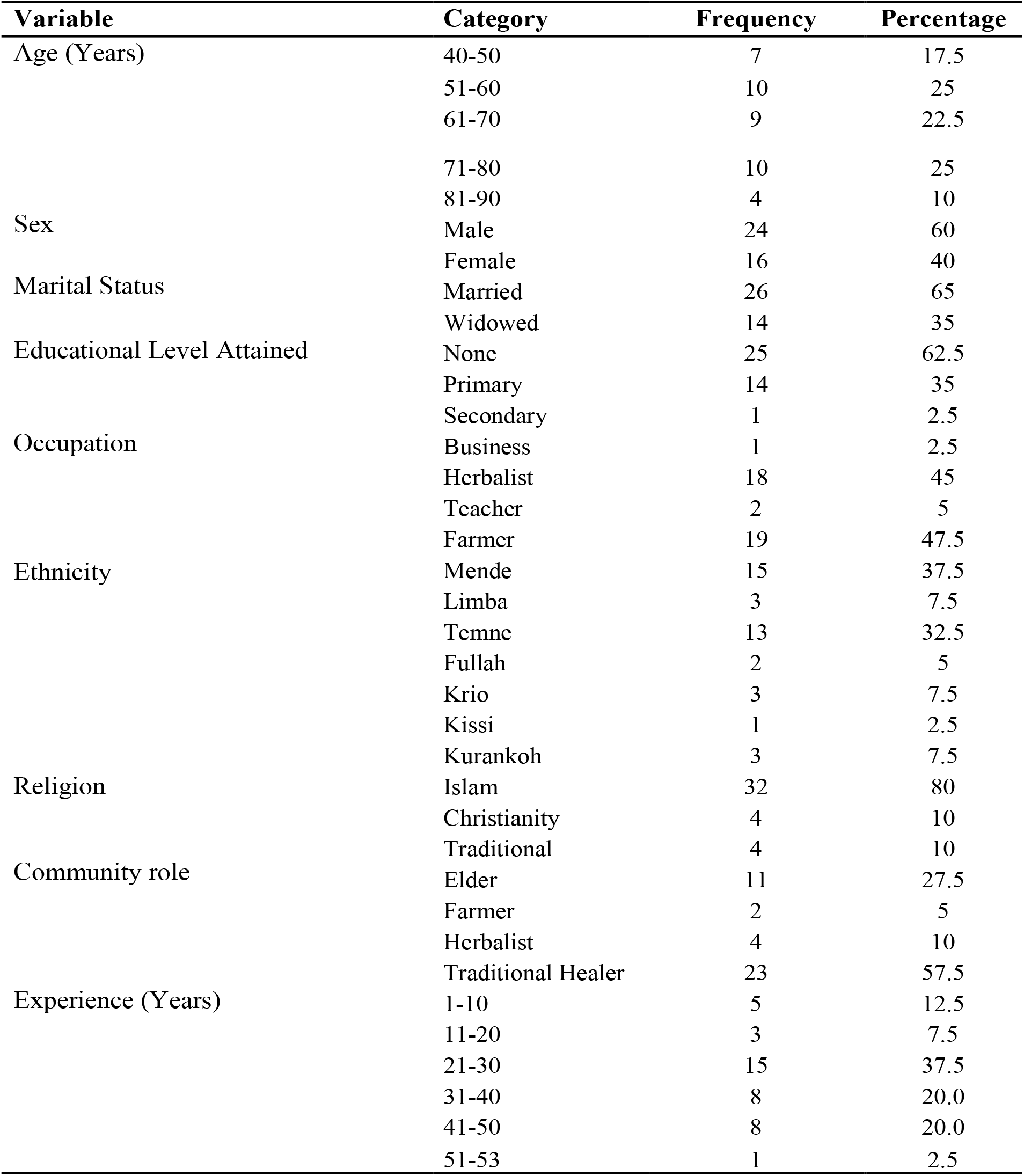
Sociodemographic Characteristics of Study Participant (N=40)

#### Diversity of Medicinal Plants

A total of 21 medicinal plant species belonging to multiple botanical families were documented.

### 3.2 Frequency and Cultural Importance

*Moringa oleifera* was the most cited species (FC = 9; RFC = 0.225), followed by:

- *Vernonia amygdalina* (FC = 7)
- *Cassia siberiana* (FC = 6)
- *Telfairia occidentalis* (FC = 6)

**Figure 1.**
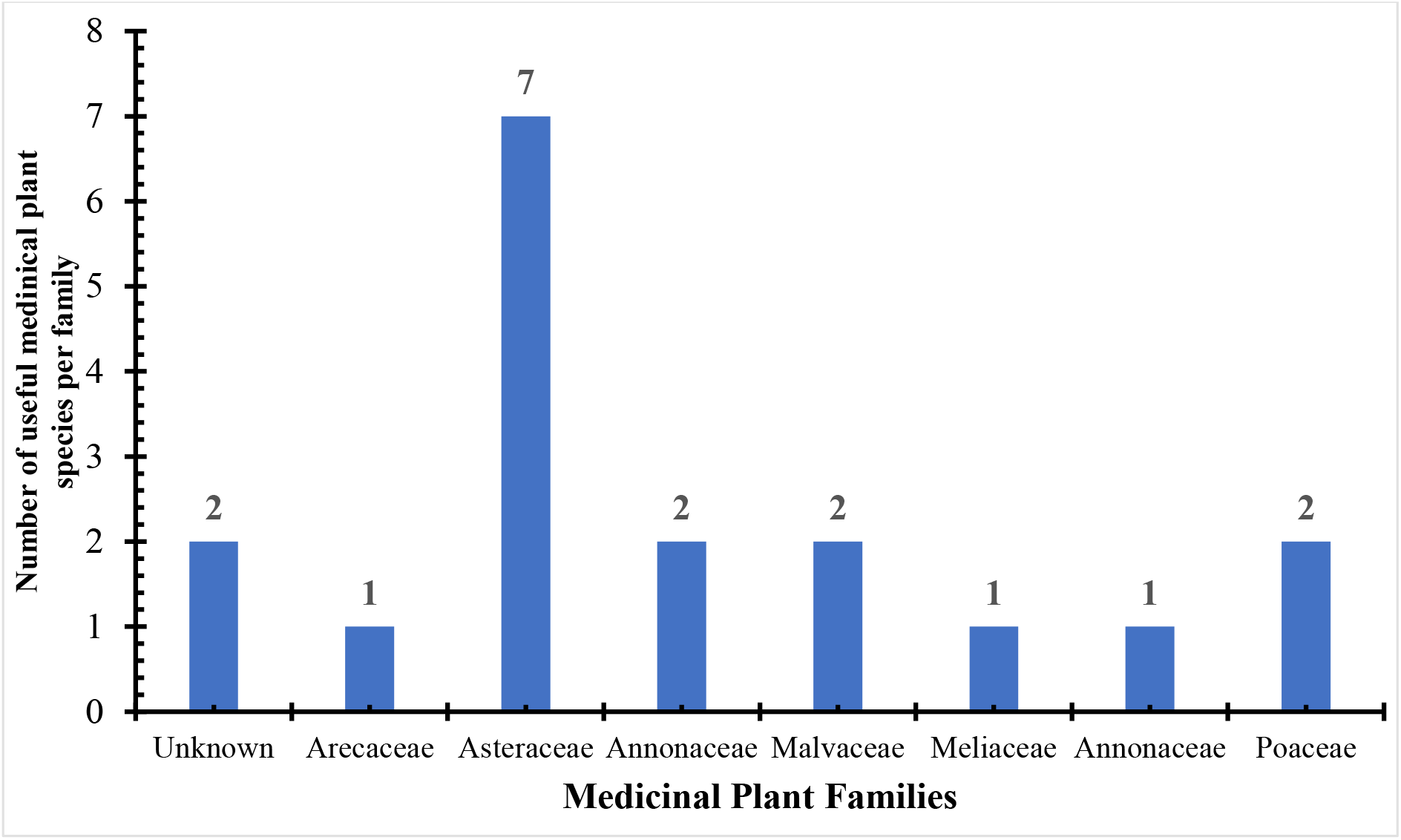
Number of useful medicinal plant species per family.

### 3.3 Informant Consensus

The calculated ICF (0.69) indicates strong agreement among informants.

### 3.4 Plant Parts Used

- Leaves: 40.9%
- Roots: 18.1%
- Bark: 13.6%

**Figure 2.**
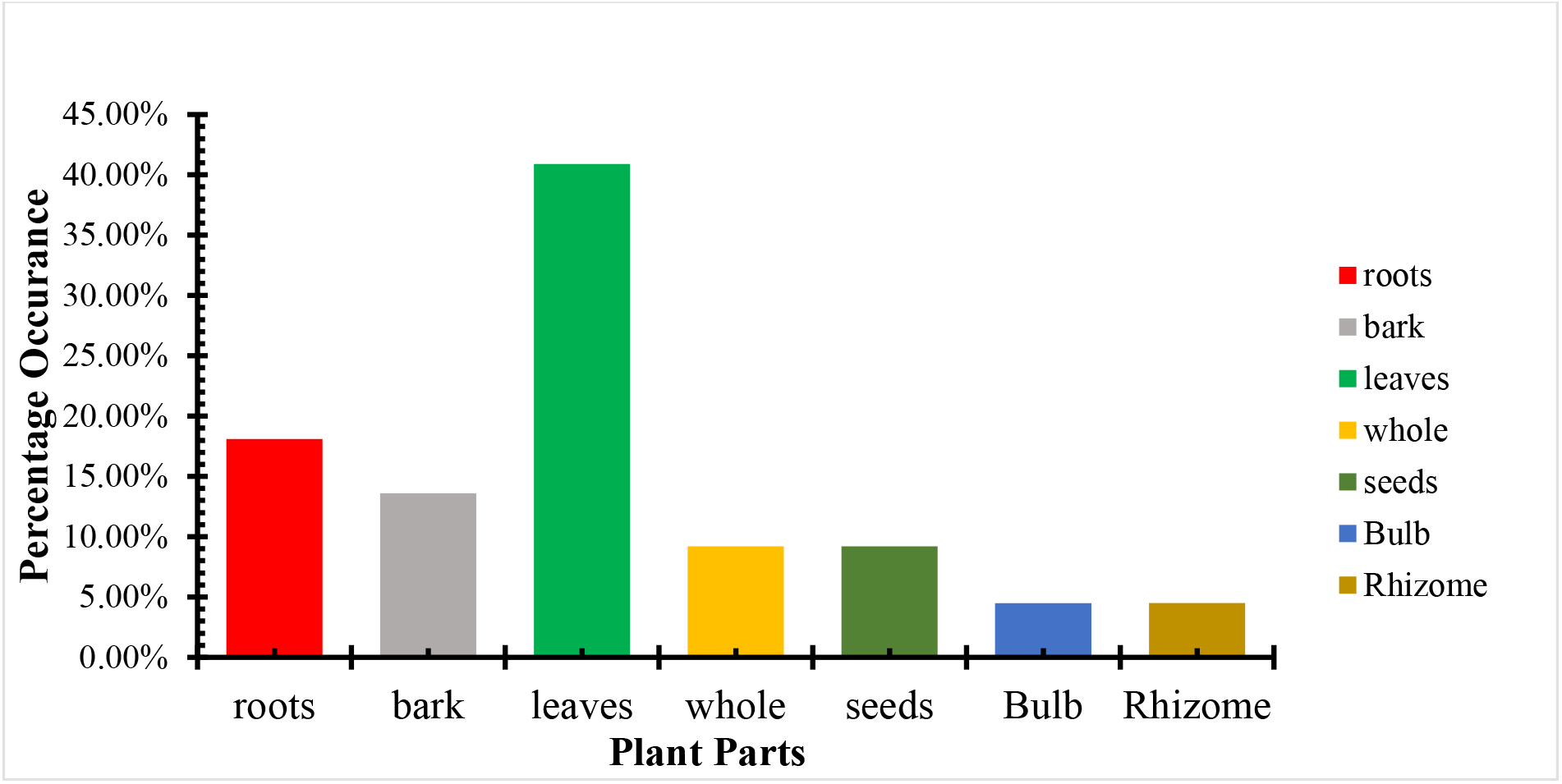
Patterns of Plant Part utilization.

### 3.5 Preparation Methods

- Decoction: 76.2%
- Powder: 14.3%
- Others: 9.5%

### 3.6 Routes of Administration

- Oral: 95.2%
- Topical: 4.8%

**Table 2.**
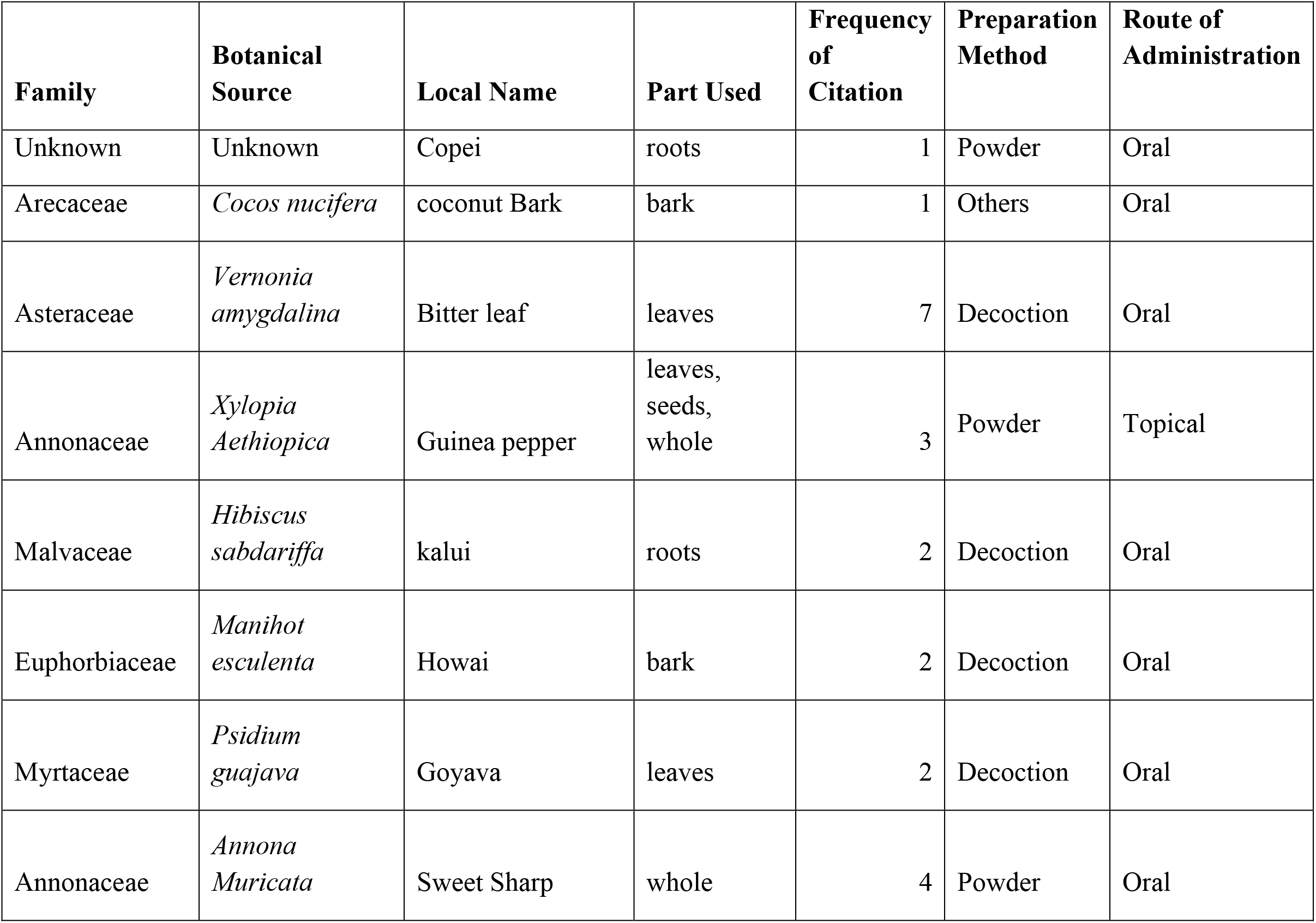
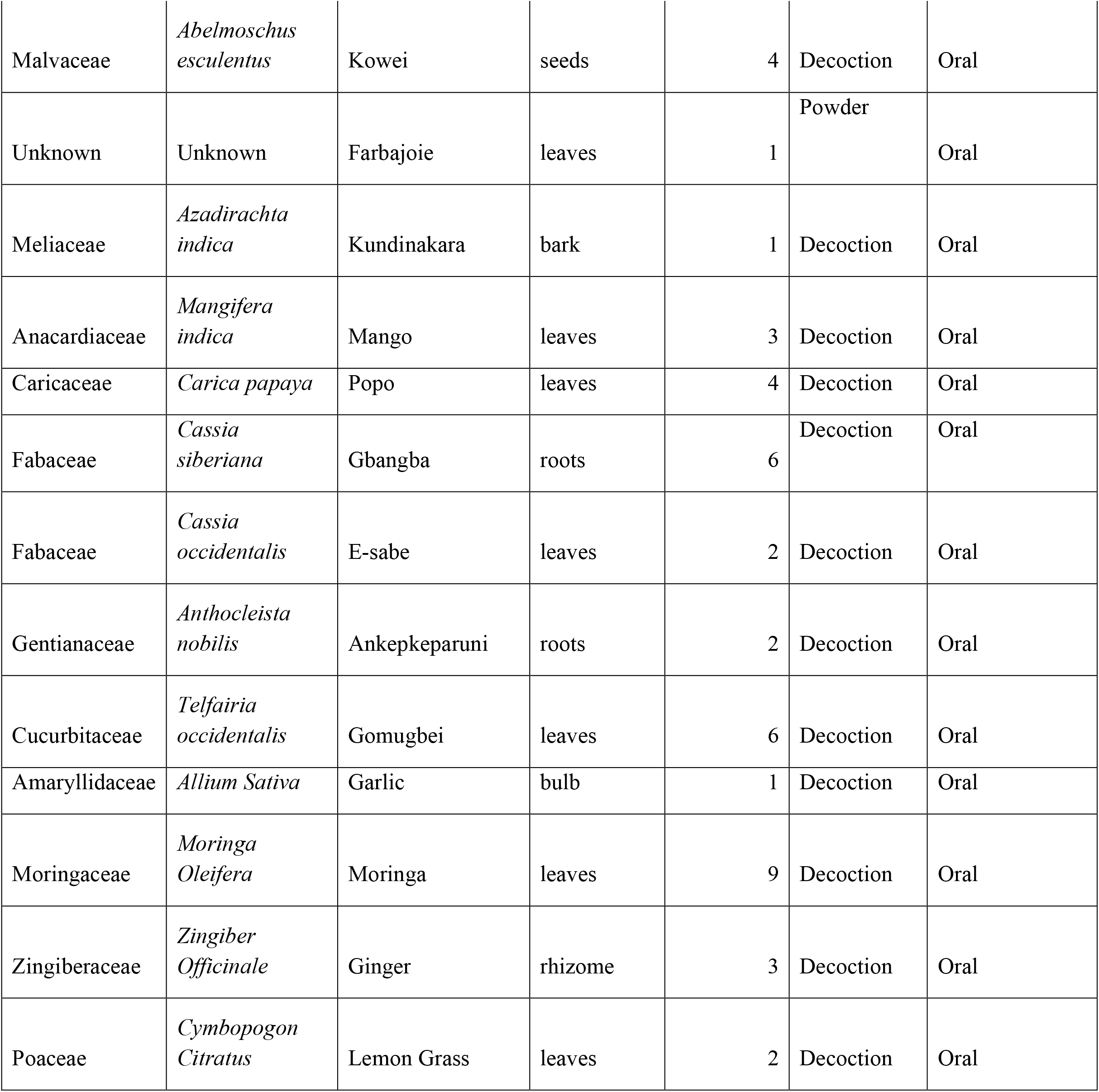
Frequently cited medicinal plant species used in diabetes management showing plant parts, preparation methods, and routes of administration.

### 3.7 Knowledge Transmission

- Apprenticeship: 37.5%
- Inheritance: 37.5%
- Experience: 25%

**Figure 3.**
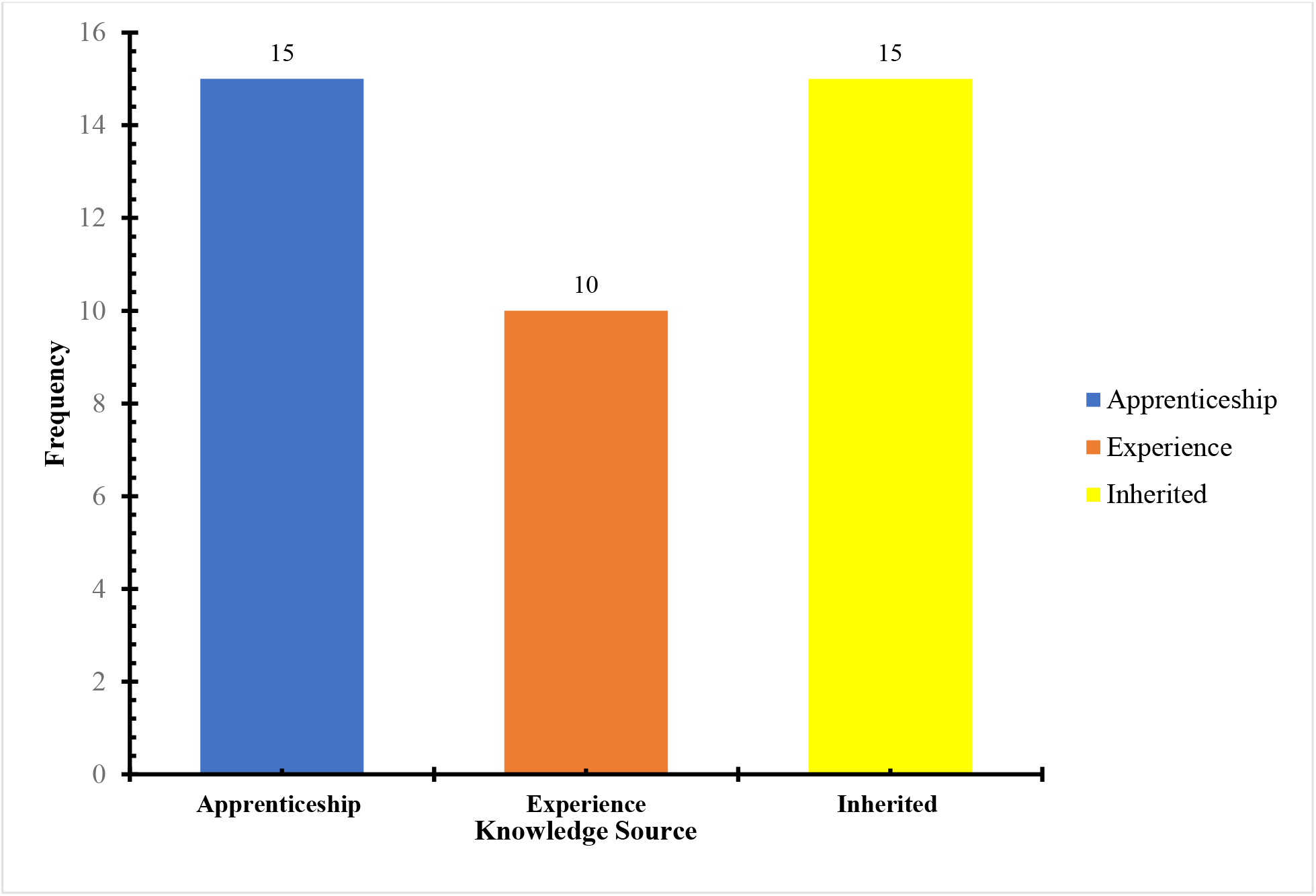
Knowledge Transmission Pathways.

## 4. Discussion

This study highlights the continued importance of medicinal plants in diabetes management in Sierra Leone. The identification of 21 plant species reflects a rich ethnobotanical knowledge system within the study communities.

The findings of this study demonstrate that traditional medicinal knowledge related to diabetes management is predominantly transmitted through apprenticeship and inherited family knowledge. This pattern is consistent with ethnobotanical studies conducted in Ghana, Ethiopia, and South Africa, where knowledge transfer occurs mainly through oral traditions and mentorship systems (Wodah & Asase, 2012; Agize et al., 2022; Constant & Tshisikhawe, 2018). Similar patterns of knowledge transmission have been reported in other African contexts, where traditional ecological knowledge is maintained through oral systems and community practices (Muvengwi & Maroyi, 2025).

The prominence of *Moringa oleifera* is supported by its well-documented phytochemical and pharmacological properties, including antioxidant and hypoglycemic effects (Leone et al., 2015; Abd Rani et al., 2018). The presence of plant species such as *Vernonia amygdalina* in this study aligns with findings from other African ethnobotanical surveys. For instance, studies in Uganda and Nigeria have reported these species among the most frequently used plants for diabetes management, largely due to their documented hypoglycemic and antioxidant properties (Tugume & Nyakoojo, 2019; Alam et al., 2022). The repeated citation of these plants across different geographical regions suggests a strong ethnopharmacological consensus and supports their potential for further scientific validation.

The predominance of leaves as the most commonly used plant part is also consistent with previous studies, where leaves are preferred due to their accessibility, ease of harvesting, and high concentration of bioactive compounds (Amuri et al., 2018). However, the use of roots and bark observed in this study raises sustainability concerns, as these practices may lead to overharvesting and depletion of plant species if not properly managed.

The dominance of decoction as the primary preparation method reflects a widespread traditional practice in African herbal medicine. Decoction facilitates the extraction of water-soluble phytochemicals, which are often responsible for therapeutic effects (Chahrour et al., 2025). Similar findings have been reported in ethnobotanical studies in Ethiopia and the Democratic Republic of Congo, where decoction is the most commonly used preparation method for antidiabetic remedies (Agize et al., 2022; Amuri et al., 2018).

The high Informant Consensus Factor (ICF) observed in this study suggests a strong level of agreement among respondents regarding the use of specific plant species for diabetes management. High consensus values are often interpreted as indicators of well-established and culturally validated medicinal practices (Ong et al., 2020). This reinforces the reliability of the ethnobotanical knowledge documented in this study.

Taken together, the findings of this study demonstrate that medicinal plant use for diabetes management in Sierra Leone is not arbitrary but reflects a structured system of knowledge supported by shared community practices. The high level of informant consensus and repeated citation of specific plant species indicate culturally validated therapeutic approaches that have been refined over time. These patterns, combined with the dominance of specific plant parts and preparation methods, provide important insights into how traditional medicine is practiced and sustained. These observations form a strong basis for further scientific investigation and underscore the relevance of ethnobotanical knowledge in addressing contemporary health challenges such as diabetes mellitus.

## 5. Conclusion

This study demonstrates that traditional medicinal plants remain an important component of diabetes management in Sierra Leone, particularly in communities where access to conventional healthcare services is limited. A total of twenty-one medicinal plant species were documented, reflecting a rich diversity of ethnobotanical knowledge. Among these, *Moringa oleifera, Vernonia amygdalina, Cassia siberiana*, and *Telfairia occidentalis* emerged as the most frequently cited and culturally significant species, indicating their prominent role in local therapeutic practices.

The predominance of leaves as the most commonly utilized plant part, along with the widespread use of decoction as the primary preparation method and oral administration as the main route of treatment, highlights consistent patterns in traditional remedy formulation. The relatively high Informant Consensus Factor further suggests that the selection and use of these medicinal plants are based on shared, structured, and culturally validated knowledge rather than random practice.

Importantly, the study underscores the central role of apprenticeship and inherited knowledge in the transmission of ethnomedicinal practices, emphasizing the dependence of traditional medicine on oral and intergenerational learning systems. However, this reliance also presents a significant risk of knowledge erosion due to ongoing socio-cultural changes, urbanization, and declining interest among younger generations.

Overall, the findings of this study provide a valuable baseline for future research, particularly in the areas of phytochemical characterization, pharmacological validation, and safety assessment of the most frequently cited plant species. In addition, the study highlights the need for systematic documentation and preservation of indigenous knowledge, as well as the promotion of sustainable harvesting practices to ensure long-term availability of medicinal plant resources. Integrating scientifically validated traditional remedies into national healthcare frameworks may also offer an opportunity to improve access to affordable and culturally acceptable diabetes management options in Sierra Leone.

Future interdisciplinary collaboration between researchers, traditional healers, and policymakers will be essential in translating ethnobotanical knowledge into evidence-based therapeutic applications.

## 6. Recommendations

Based on the findings of this study, the following recommendations are proposed:

- **Phytochemical and pharmacological validation:** Further laboratory-based studies should be conducted on the most frequently cited plant species, particularly *Moringa oleifera, Vernonia amygdalina, Cassia siberiana*, and *Telfairia occidentalis*, to isolate active compounds, evaluate mechanisms of action, and establish their efficacy and safety profiles in the management of diabetes mellitus.
- **Toxicological and dosage standardization studies:** There is a need for controlled toxicological assessments and dosage standardization of commonly used herbal preparations to ensure safe use and minimize the risk of adverse effects associated with unregulated consumption.
- **Sustainable harvesting and conservation strategies:** Given the use of roots and bark in some remedies, which may threaten plant survival, conservation measures should be implemented. These include promoting the use of renewable plant parts (such as leaves), encouraging cultivation of medicinal plants, and integrating community-based conservation practices.
- **Documentation and preservation of indigenous knowledge:** Systematic documentation of ethnobotanical knowledge should be prioritized to prevent its loss due to urbanization, generational shifts, and declining interest among younger populations. Digital archiving and community-based knowledge repositories may serve as effective preservation strategies.
- **Integration into healthcare systems:** Policymakers should explore pathways for integrating scientifically validated traditional remedies into national healthcare frameworks as complementary approaches to diabetes management, particularly in underserved communities.
- **Capacity building and community education:** Training programs should be developed for traditional healers and community members on safe preparation methods, hygiene, dosage practices, and recognition of complications requiring referral to formal healthcare facilities.
- **Interdisciplinary collaboration:** Strong collaboration between researchers, healthcare professionals, traditional healers, and regulatory authorities should be encouraged to facilitate knowledge exchange, improve research translation, and support evidence-based use of medicinal plants.
- **Further ethnobotanical surveys:** Additional large-scale studies across other regions of Sierra Leone are recommended to capture a more comprehensive inventory of medicinal plants used in diabetes management and to validate the patterns observed in this study.

These recommendations provide a framework for translating ethnobotanical knowledge into evidence-based interventions for diabetes management.

## Data Availability

All data produced in the present study are available upon reasonable request to the authors

